# Examining hearing-aid benefit during story listening in older adults using large language models

**DOI:** 10.64898/2026.10.22.26366255

**Authors:** Yue Ren, Noble Iwuoha, Sarah Bobbitt, Tara Raessi, Sylvia Ciechanowski, Karen A. Gordon, Björn Herrmann

## Abstract

Hearing-aid benefit is typically assessed with isolated sentences, whereas everyday listening involves continuous speech. Yet, scoring comprehension of such speech can be labor-intensive. Here, we examined whether large language models (LLMs) can assess speech intelligibility and story comprehension in older hearing-aid users, and identify who benefits from hearing aids. Thirty-nine older adults (median age: 75 years) listened to sentences and short stories in multi-talker babble, under aided and unaided conditions. LLM-based intelligibility scores were nearly identical to established, rule-based scores. LLM-based intelligibility, gist, and detail recall scores were sensitive to masking and hearing-aid use. On average, hearing aids improved sentence intelligibility and story recall. Benefits increased with poorer hearing and, independently, with greater audibility gains from hearing aids. However, about half of the participants showed little benefit, primarily those with milder (though still clinically significant) hearing loss or smaller improvements in audibility. The results demonstrate that benefits of hearing aids for sentence intelligibility extended to comprehension of short stories and that LLMs enable automated assessment of both.

## Introduction

Hearing loss is highly prevalent in older adulthood (Goman and Lin, 2016; Feder et al., 2017) and is associated with communication difficulties, social isolation, and cognitive decline (Nachtegaal et al., 2009; Livingston et al., 2024). Hearing aids are the primary intervention for age-related hearing loss. Many hearing-aid users report benefit and satisfaction (Picou, 2020, 2022), but benefit varies widely across individuals, and remaining difficulties understanding speech in noise are a common reason for dissatisfaction with and non-use of hearing aids (Knudsen et al., 2010; McCormack and Fortnum, 2013; among other factors: Bainbridge and Wallhagen, 2014; Cumming et al., 2026). Identifying who benefits from hearing aids and in what types of listening environments remains unclear.

Hearing aids primarily restore audibility through frequency-specific amplification. Restored audibility improves speech perception relative to unaided listening (Ferguson et al., 2026), particularly for soft and conversational-level speech in quiet (Larson et al., 2000; Shanks et al., 2002; Kuk et al., 2015). In background noise, aided benefit to speech perception is typically more limited (Moore et al., 1985; Larson et al., 2000; Shanks et al., 2002; Kuk et al., 2015; Löhler et al., 2015). This may be partly because amplification does not reverse central auditory changes that develop with untreated hearing loss, such as increased neural excitability and internal neural noise (Auerbach et al., 2014; Herrmann and Butler, 2021), or compensate for cognitive limitations (Humes, 2007; Lunner et al., 2009) and suprathreshold deficits, such as reduced frequency selectivity and temporal processing (Souza, 2016; Lesica, 2018). Benefit can be limited further because clinical fittings might not fully restore audibility (Humes, 2002, 2007; Lopez-Poveda et al., 2017) and because compression, which reduces gain at higher input levels, can alter the effective signal-to-noise ratio (SNR; Souza, 2002; Souza, 2016). Consequently, hearing-aid benefit for speech perception differs across listeners, and which listeners benefit most remains difficult to predict from the audiogram alone (e.g., using the Speech Intelligibility Index; Ching et al., 1998; Humes, 2002).

Speech-in-noise perception is typically assessed by asking listeners to repeat short, unrelated sentences verbatim (e.g., HINT, QuickSIN; Nilsson et al., 1994; Killion et al., 2004). Some tests present sentences within thematically related passages, such as the Connected Speech Test (Cox et al., 1987; Cox et al., 1988), which has been used to quantify hearing-aid benefit (Cox and Alexander, 1992; Larson et al., 2000; Humes, 2002), but these tests still score verbatim, sentence-by-sentence repetition of key words rather than what listeners understood or retained from the passage as a whole. Everyday listening, however, often involves continuous speech in which a topic unfolds over tens of seconds (Eisenberg, 1985; Ochs et al., 1992; Bohanek et al., 2009) in background noise at moderately positive SNRs (Smeds et al., 2015; Wu et al., 2018). Individuals typically retain the gist of speech rather than its exact wording (Mehler and Miller, 1964; Sachs, 1967; Schneider et al., 2002; Raccah et al., 2024), and they can become absorbed in stories (Herrmann and Johnsrude, 2020) and segment speech into larger meaningful units (Herrmann et al., 2026; Panela et al., 2026) despite moderate noise and imperfect intelligibility. Findings for disconnected sentences may thus not generalize to continuous speech (Keidser et al., 2020), such that age-related intelligibility observations do not always translate from sentence to story listening (Irsik et al., 2022). Moreover, being able to recall and retell what was heard – both its gist (the main ideas and overall meaning) and its specific details – may also be an important component of successful listening, one that verbatim sentence report does not assess, because retelling is rarely verbatim. It remains unclear whether hearing aids improve comprehension of continuous speech, such as stories, and which individuals benefit most.

Assessing the comprehension of continuous speech has been limited by practical constraints. Naturalistic paradigms typically require writing story texts, recording them with voice actors, and manually transcribing and scoring open-ended free recall by trained raters, all of which is labor-intensive (Ward et al., 2016; Martinez, 2024; van Genugten and Schacter, 2024). Large language models (LLMs) may overcome these constraints. They can generate stories of controlled length and content (Herrmann, 2025d, b; Lamekina et al., 2026), in different languages (Herrmann, 2025d; Polspoel et al., 2025), synthesize naturalistic speech (Shen et al., 2017; Herrmann, 2023; Bruder et al., 2025; Herrmann, 2025d; Lavan et al., 2025), and transcribe spoken responses (Radford et al., 2022). Verbatim intelligibility can be scored automatically using rule-based human-like approaches (Borrie et al., 2019; Stipancic et al., 2025) or LLMs (Herrmann, 2025c; Zhang et al., 2025). LLMs and related approaches can further quantify how well a person’s recall captures the meaning of a story, either through the semantic similarity of text embeddings (numerical vectors capturing semantic content of speech) or through prompted ratings (Heusser et al., 2021; Raccah et al., 2024; Herrmann, 2025d; Panela et al., 2026), and can identify which story details were recalled (Martinez, 2024; Georgiou et al., 2025). Such scores agree well with human raters (Chandler et al., 2021; Martinez, 2024; Panela et al., 2025) and are sensitive to background noise (Herrmann, 2025d; Panela et al., 2026). However, these approaches have mainly been tested in younger adults with normal hearing. Whether they are equally sensitive in older adults with hearing loss and to aided versus unaided listening is unknown. Moreover, most work has used proprietary LLMs, which require uploading participant data to company servers, raising privacy concerns in clinical use that locally run open-source models could avoid.

The current study had two aims. First, we examined whether LLMs, including locally run open-source models, can assess speech intelligibility and story comprehension in older adults who use hearing aids. Second, we investigated whether hearing aids benefit story comprehension as they benefit sentence intelligibility, and whether benefit can be predicted using pure-tone average (PTA) and the audibility benefit individuals obtain from their hearing aids. We expected: 1) LLM-based scores to closely match established intelligibility scoring; 2) hearing-aid benefit to be greater for individuals with significant hearing loss and larger gains in audibility from hearing aids; and 3) gist recall to be less affected than intelligibility by masking and hearing-aid use.

## Methods and materials

### Participants

Thirty-nine hearing-aid users took part (median age: 75 years; range: 50–88 years; 16 male, 23 female). All were native English speakers or had learned English before age 5. Participants had at least 6 months of hearing-aid experience (median: 7 years; mean: 9.8 years; range: 0.5–37 years), had used their current aids for 1 month to 8 years (median: 3 years; mean: 2.7 years), and wore them for 1–16 hours daily (median: 11 hours; mean: 10.1 hours). They used their own behind-the-ear (n = 35) or in-the-ear (n = 3) aids from different manufacturers (not recorded for one person). No participant reported a neurological disease, except one with trigeminal neuralgia, which did not affect participation. Four additional participants were excluded: Three did not complete the main parts of the study (two withdrew due to task difficulty or fatigue; one could not perform the tasks due to aphasia), and one data set was accidentally overwritten. Participants gave written informed consent and were paid $10 CAD per half-hour. The study followed the Declaration of Helsinki and the Canadian Tri-Council Policy Statement (TCPS2-2022) and was approved by the Research Ethics Board of the Rotman Research Institute at the Baycrest Academy for Research and Education.

### Experimental setup

Participants sat in a single-wall sound booth (Eckel) facing a ViewSonic monitor (60 Hz) that mirrored the experimental computer (Windows 10; Psychtoolbox v3.0.14 in MATLAB, MathWorks Inc.). Sounds were presented via an RME Fireface UCX external sound card: speech at 65 dB SPL from a horizontal speaker (SADA V-196, SADA Technology) in front of the participant (∼0.8 m, aligned with the bottom of the screen), and background sound at different signal-to-noise ratios (SNRs) from four speakers (JBL Control 1 Pro) placed at 45-degree angles (left front, right front, left behind, right behind) at a distance of ∼1.2 m from the participant’s head. Verbal responses were recorded with a Shure SM7B microphone.

### Hearing assessment

Unaided air-conduction pure-tone audiometry was administered at 0.25, 0.5, 1, 2, 4, and 8 kHz, and thresholds were averaged across ears. The PTA was calculated as the average across 0.5, 1, 2, and 4 kHz (Stevens et al., 2013; Humes, 2019).

### Stimulus materials, procedures, and analyses

Throughout the study, aided listening (hearing aids at the setting participants commonly use in speech-in-noise contexts) was contrasted with unaided listening (hearing aids removed). The following three sections describe the protocols in the order in which they were administered. Data were analyzed in Python, R (v4.5.2; Autoscore; Borrie et al., 2019), and MATLAB, and statistical analyses were carried out in JASP (v0.19.1.0) and MATLAB.

#### Estimation of sensation level and audibility benefit

To quantify how much hearing aids improved audibility for each participant, we measured an analogue of functional gain, the difference between unaided and aided sound-field thresholds traditionally used to verify hearing-aid fittings (Haskell, 1987; Humes and Kirn, 1990; Kuk and Ludvigsen, 2003). The sensation level for pink noise was estimated using a method-of-limits procedure, separately for aided and unaided listening (Leek, 2001; Herrmann and Johnsrude, 2018; Herrmann et al., 2022). Pink noise was used instead of frequency-specific tones because it spans the speech frequency range and, like speech, has more energy at low than at high frequencies, and because a single broadband measure avoided lengthy frequency-by-frequency testing. A 15-s pink noise continuously decreased (from suprathreshold levels) or increased (from subthreshold levels) in intensity at 5 dB/s, and participants pressed a button, which stopped the sound, when they could no longer hear or started to hear the noise, respectively. The intensity at the button press was averaged across 8 decreasing and 8 increasing sounds (alternating) to obtain the sensation level. The order of aided and unaided listening was counterbalanced across participants. Audibility benefit was calculated as the difference in sensation level between unaided and aided listening. Because hearing aids apply level-dependent (compressive) gain, this threshold-based measure reflects the audibility of soft sounds, whereas the gain for suprathreshold levels, such as conversational speech, may be smaller (Kuk and Ludvigsen, 2003). Audibility benefit was therefore treated as an index of individual differences in audibility rather than as an estimate of the gain applied to speech. It was used as a continuous predictor of speech-perception outcomes, and a median split (2.59 dB) was used for some visualizations.

#### Intelligibility of disconnected sentences in multi-talker babble

For the sentence-intelligibility task, sixty 8-word sentences were created iteratively using OpenAI’s GPT-4o mini (OpenAI et al., 2023) with manual verification (e.g., ‘Rescue arrived just as his strength began fading.’). Modern large language models can generate synthesized speech that mirrors human production properties (Lamekina et al., 2026) and is increasingly indistinguishable from human speech (Herrmann, 2023; Bruder et al., 2025; Herrmann, 2025d; Herrmann and Cui, 2025; Lavan et al., 2025). Auditory sentences were thus synthesized using OpenAI’s text-to-speech model (‘tts-1-hd’, ‘Alloy’ voice) via OpenAI’s application programming interface (API), with durations ranging from 2.31 to 4.02 s (mean: 3.05 s).

Sentences were presented from the front speaker, and twelve-talker babble (Bilger, 1984; Bilger et al., 1984; Wilson et al., 2012) was played concurrently from the four surrounding speakers (same signal), starting 2.5 s before sentence onset (enabling hearing-aid adjustment) and ending 0.4 s after sentence offset. Three sentences were presented at each of 10 equally spaced SNRs (–5 to +20 dB), manipulated by changing the babble level while speech was kept constant at 65 dB SPL. This corresponds to a typical conversational level (Pearsons et al., 1977; Sato et al., 2011; Šrámková et al., 2015) and falls within the range where hearing-aid intelligibility benefits have been observed (Larson et al., 2000). Assignment of sentences to SNRs was randomized. Participants completed the task twice (aided and unaided; order counterbalanced), with 30 different sentences each, and verbally repeated each sentence exactly as they heard it.

To determine the SNRs for the story-listening task, aided responses were analyzed in real time. Responses were transcribed using OpenAI’s ‘gpt-4o-transcribe’ model (Python API; language set to English), and the proportion of correctly reported words was calculated using Autoscore, an R library whose estimates correlate highly with manually scored intelligibility (Borrie et al., 2019; Herrmann, 2025c; Stipancic et al., 2025). A logistic function was fit to the proportion of correctly reported words across the 10 SNRs:

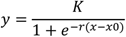

where *y* is the proportion of correctly reported words, *x* the SNRs, *K* the upper asymptote (maximum proportion correct), *r* the slope, and *x0* the SNR at the function’s midpoint (inflection point). *K* was fixed at 1, whereas *r* and *x0* were free. From the fit, the SNRs yielding 75% and 95% correctly reported words under aided listening (SNR75 and SNR95, respectively) were calculated and used, together with speech in quiet, in the story-listening task. Mean SNR75 and SNR95 were 8.8±6.8 dB and 18.5±10.5 dB, respectively (±standard deviation).

For the offline analysis, the proportion of correctly reported words was calculated for each SNR using Autoscore, separately for aided and unaided listening. Previous work has also shown that large language models can score speech intelligibility and comprehension (Herrmann, 2025c, d; Panela et al., 2025; Panela et al., 2026), but this has not been tested in older adult hearing-aid users. Therefore, we also scored intelligibility using an LLM (‘gpt-5-mini-2025-08-07’), which was given numbered word lists of the target sentence and the response transcript and instructed to apply Autoscore’s scoring rules (e.g., word order ignored, homophones counted as correct, morphological changes and synonyms counted as incorrect). Because LLM outputs can vary across calls, all LLM-based analyses were repeated five times per response, and scores were averaged across repetitions. The exact prompt is provided in the Supplementary Materials. The proportion of correctly reported words was calculated for each SNR, separately for aided and unaided listening.

For both Autoscore and the LLM, a logistic function was fit with all three parameters free, because the upper asymptote (*K*) varied strongly across participants and conditions. Estimated parameters were analyzed using a repeated-measures analysis of variance (rmANOVA) with the within-participant factors Device Use (aided, unaided) and Analysis Approach (Autoscore, LLM). PTA, audibility benefit, and age were subsequently added as covariates. Main effects and interactions of within-participant factors are reported from models without covariates. Models with covariates were used only to test interactions between covariates and Device Use; i.e., associations with the hearing-aid benefit. Agreement between Autoscore and LLM scores was quantified using intraclass correlations (ICC; two-way model, absolute agreement, single measures; McGraw and Wong, 1996; Koo and Li, 2016).

#### Story listening in multi-talker babble

In the main task, participants listened to stories in 12-talker babble. Eighteen story texts on different topics (e.g., a rivalry that turns into respect) were generated (Georgiou et al., 2025; Herrmann, 2025a) with OpenAI’s GPT-4o (’gpt-4o-2024-08-06’; OpenAI et al., 2023), using a zero-shot prompt (Chen et al., 2025) that requested a single-paragraph story with simple vocabulary and a final sentence whose meaning depends on the preceding context (prompts and more details are in the Supplementary Materials). Stories comprised 118–121 words and were synthesized using OpenAI’s Alloy voice (’tts-1-hd’; ∼44–50 s). The stories’ final sentences (eight words, seven for two stories; ∼2.5–3.7 s) were used to assess intelligibility.

Stories were presented in quiet or at the participant’s aided SNR75 or SNR95, with speech and babble presented as in the sentence task, in a fully crossed 2 × 3 design (Device Use: aided, unaided; Speech Clarity: SNR75, SNR95, in quiet; three randomly assigned stories per condition). Stories were presented in 6 blocks, each comprising the three speech-clarity conditions in randomized order. Aided and unaided blocks alternated, with the starting condition counterbalanced across participants. After each story, participants verbally repeated its last sentence exactly as they heard it (intelligibility) and then recalled the entire story in as much detail as possible (comprehension). Both responses were transcribed using OpenAI’s ‘gpt-4o-transcribe’ model. Two practice stories familiarized participants with the task.

#### Analysis of sentence intelligibility during story listening

The proportion of correctly reported words of the final sentence was calculated using Autoscore (Borrie et al., 2019) and the LLM (as for the sentence-in-noise task), separately for each speech-clarity and hearing-aid condition.

An rmANOVA was calculated on the proportion of correctly reported words, with Device Use (aided, unaided), Speech Clarity (SNR75, SNR95, in quiet), and Analysis Approach (Autoscore, LLM) as within-participant factors. PTA, audibility benefit, and age were subsequently added as covariates to examine their association with the intelligibility benefit from hearing aids. Post hoc tests were corrected for multiple comparisons using Holm’s method (Holm, 1979).

#### Analysis of story comprehension

Story comprehension was assessed by scoring the gist and the recalled details of each recall transcript relative to the original story text. Autoscore is unsuitable here, because it compares responses verbatim with the original speech, whereas story recall is not verbatim but reflects the gist of what was heard. Large language models can automate comprehension scoring, and LLM scores correlate well with human ratings (Martinez, 2024; Georgiou et al., 2025; Panela et al., 2025).

For gist, the LLM (OpenAI’s ‘gpt-5-mini-2025-08-07’) was provided with the story text and the recall transcript and prompted to ‘rate the degree to which the gist of the story was recalled by the participant’ on a scale from 0 (‘not very’) to 100 (‘very’), using ‘the full range of the scale’ (Herrmann, 2025d; Panela et al., 2025; full prompt in the Supplementary Materials). Ratings were divided by 100 (gist score from 0 to 1) and averaged across the five repetitions and the three stories per condition.

For details, recall was scored in two steps using the LLM, following recent automated approaches to memory detail scoring (Martinez, 2024; van Genugten and Schacter, 2024; Georgiou et al., 2025). First, each original story was decomposed into ‘atomic recallable details’, defined in the prompt as ‘a single fact, event, state, relationship, or outcome that a participant could recall independently’, with each detail expressing ‘only ONE idea’, akin to the propositions (Kintsch and van Dijk, 1978), idea units (Sisco et al., 2013), details (Sacripante et al., 2023; van Genugten and Schacter, 2024), or clauses (Shen et al., 2023; Georgiou et al., 2025) used to score text and narrative recall. This yielded on average 23 details per story (range: 19–27). Second, the LLM scored each detail as recalled (1), partly recalled (0.5), or not recalled (0) in the recall transcript, similar to manual scoring with partial credit (Power et al., 1979; Sisco et al., 2013; Martinez, 2024). The model was instructed to ‘score based on meaning, not exact wording’ and to ‘accept paraphrases’ (full prompts in the Supplementary Materials). The proportion of recalled details was calculated as the sum of scores divided by the number of details. As for all LLM-based analyses, both steps were repeated five times for each recall (with story details extracted anew each time), and the proportion of recalled details was averaged across repetitions and across the three stories per condition.

Gist scores and the proportion of recalled details were analyzed using separate rmANOVAs with Device Use and Speech Clarity as within-participant factors. As for intelligibility, PTA, audibility benefit, and age were subsequently added as covariates, and post hoc tests were Holm-corrected.

### Proprietary versus open-source models

The results reported in the main article were obtained through analyses using OpenAI models ‘gpt-4o-transcribe’ (transcription) and ‘gpt-5-mini-2025-08-07’ (analysis). These models incur (minimal) costs and require uploading data to company servers, but are fast and need no specialized hardware. However, some circumstances or laws may require stricter privacy protection. We thus repeated all analyses using open-source models run on a local computer with a dedicated graphics processing unit (GPU; NVIDIA GeForce RTX 4080): Alibaba’s Qwen3-ASR-1.7B for transcription and Google’s Gemma 4 (‘gemma-4-26b-a4b’, executed using 4-bit quantized GGUF weights) for analysis, using the same prompts and settings. Both models were run with custom Python code. Qwen3-ASR did not require the GPU, whereas Gemma 4 was run through LM Studio, requiring the GPU (v0.4.10; ‘thinking’ disabled). Models were downloaded from the Hugging Face platform (https://huggingface.co). The results, provided in the Supplementary Materials, closely mirror those obtained with OpenAI’s models, with some smaller differences that we point out in the respective results sections. Overall, the analysis suggests that local models are a viable alternative.

## Results

### Audiometry, audibility benefit, and hearing-aid usage

The median PTA was 43.13 dB HL (unaided), and all but two participants had clinically significant hearing loss (PTA > 25 dB HL; Figure 1A). Hearing aids significantly improved audibility (t_38_ = 5.279, p = 5.5 · 10^-6^, d = 0.845; Figure 1B), showing a median benefit of 2.59 dB, with a few participants gaining more than 10 dB. Age was not correlated with PTA or audibility benefit (ps > 0.13), whereas PTA and audibility benefit weakly correlated (r = 0.324, p = 0.044; Figure 1C).

**Figure 1:**
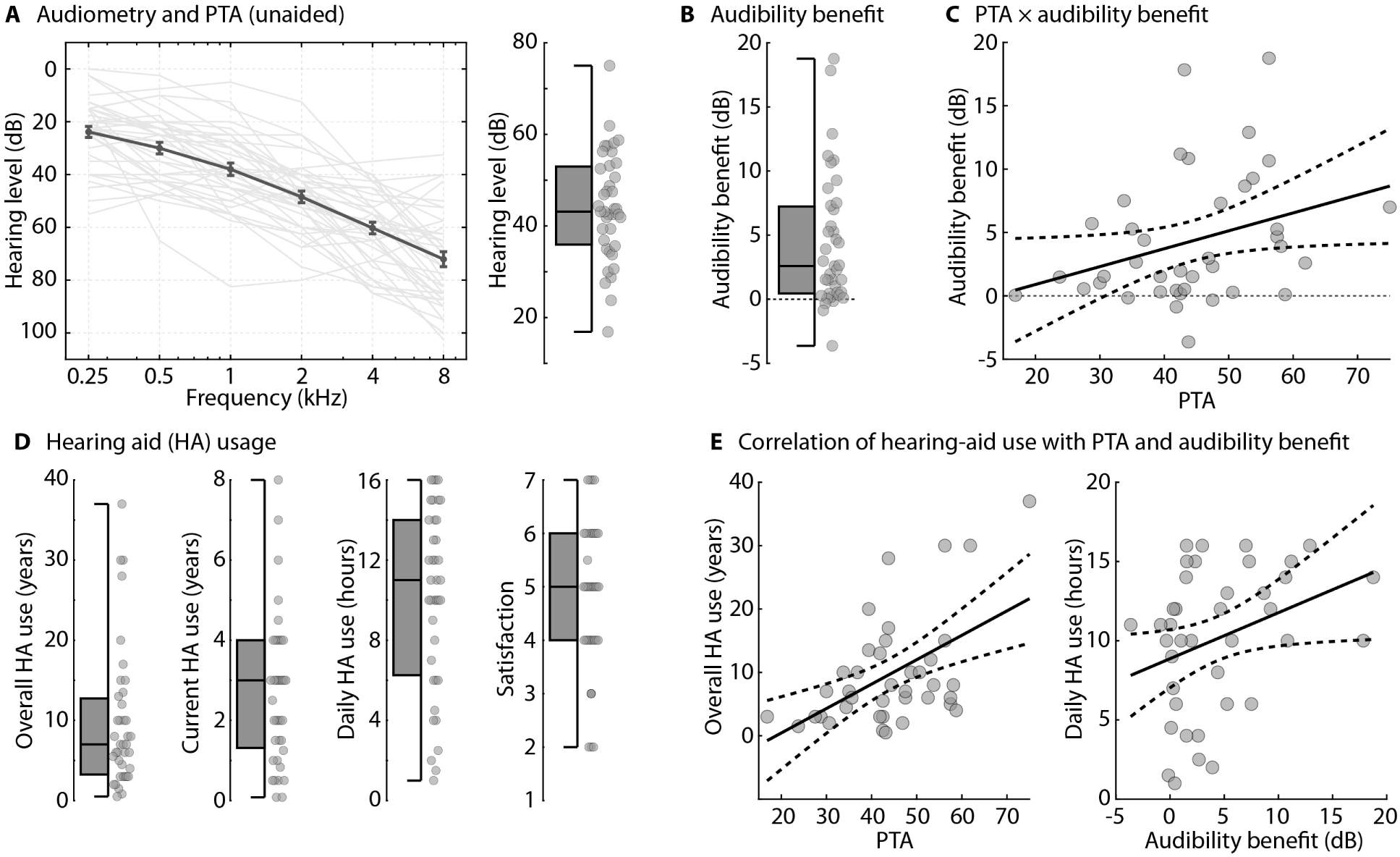
Audiometry, audibility benefit, and hearing-aid usage. **A:** Audiograms measured without hearing aids and pure-tone average threshold (PTA; average across 0.5-4 kHz). Thin lines reflect individual participants. The thick line reflects the mean (and standard error of the mean). The box plot shows the PTA, with gray dots reflecting individual participants. **B:** Hearing-aid benefit for hearing thresholds of a pink noise (unaided minus aided listening). **C:** Correlation between PTA and audibility benefit (r = 0.324, p = 0.044). **D:** Metrics of hearing-aid use: overall duration of hearing-aid use, duration of use of current hearing aids, duration of daily hearing-aid use, and satisfaction with hearing aids (1-7 scale). **E:** Relationship between PTA and overall duration of hearing-aid use (p = 0.001) and relationship between audibility benefit and duration of daily use (p = 0.047). For correlation plots, gray dots reflect individual participants, and lines reflect the best linear fit and confidence intervals.

Participants had used hearing aids for a median of 7 years (current aids: 3 years), with wide individual differences, and wore them for a median of 11 hours daily. Satisfaction was better than neutral (Figure 1D). In regression models with age, PTA, and audibility benefit as predictors, higher PTA predicted longer overall hearing-aid use (t_35_ = 3.531, p = 0.001), and larger audibility benefit predicted longer daily use (t_35_ = 2.055, p = 0.047; Figure 1E).

### Intelligibility of disconnected sentences in multi-talker babble

For the sentence-in-noise task, the upper asymptote, reflecting the performance plateau at high SNRs, was higher for aided than unaided listening (Device Use: F_1,38_ = 14.373, p = 5.2 · 10^-4^, ω^2^ = 0.092; Figure 2) and did not differ between Autoscore and LLM scoring (Analysis Approach: F_1,38_ = 0.269, p = 0.607, ω^2^ < 0.001; interaction: p > 0.6; Figures 2 and 3). The slope and midpoint of the logistic fit showed no effects or interactions (all ps > 0.09). The midpoint was slightly higher for the open-source model (Gemma 4) compared to Autoscore, suggesting slightly lower intelligibility scores (Figures S1 and S2).

**Figure 2:**
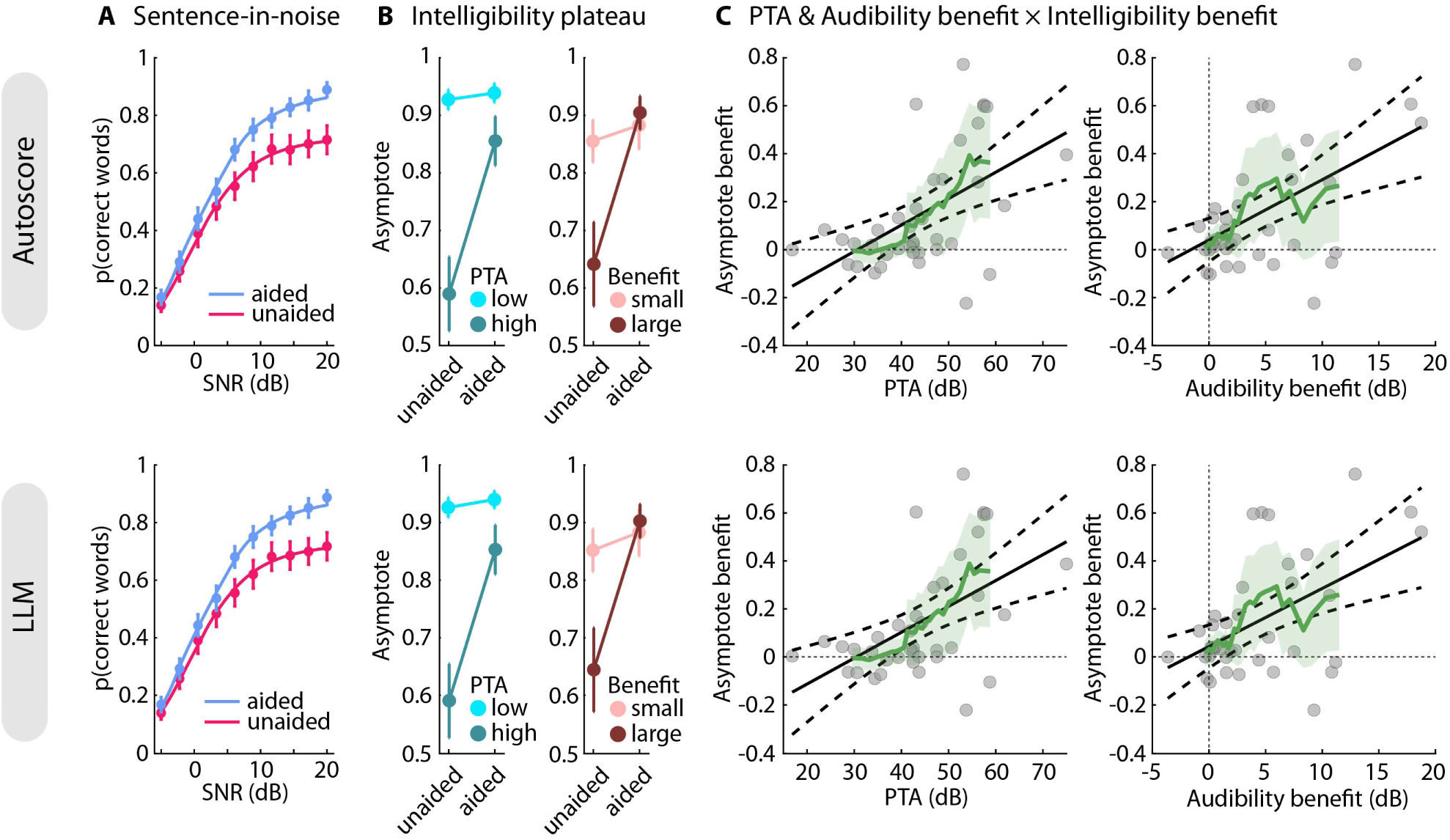
Speech intelligibility for disconnected sentences. **A:** Proportion of correctly reported words. **B:** Speech-in-noise intelligibility at the upper plateau (i.e., upper asymptote from logistic function fit), separately for participants grouped by low (16–42.5 dB HL) vs high PTA (43.13–75 dB HL) and small (-3.63–2.32 dB) vs large audibility benefit (2.59–18.76 dB) based on median split, and separately for listening with (aided) and without hearing aids (unaided). **C:** Relationship between PTA and the intelligibility benefit (i.e., upper asymptote) and the relationship between audibility benefit and the intelligibility benefit (aided minus unaided). PTA and audibility benefit both contributed independently to the intelligibility benefit. Gray dots reflect individual participants. Black lines reflect the best linear fit and 95% confidence intervals. The green line and green shaded area reflect the mean and 95% confidence interval across 10 neighboring participants, shifted along the x-axis. The top row and bottom rows show data scored using Autoscore and an LLM, respectively.

**Figure 3:**
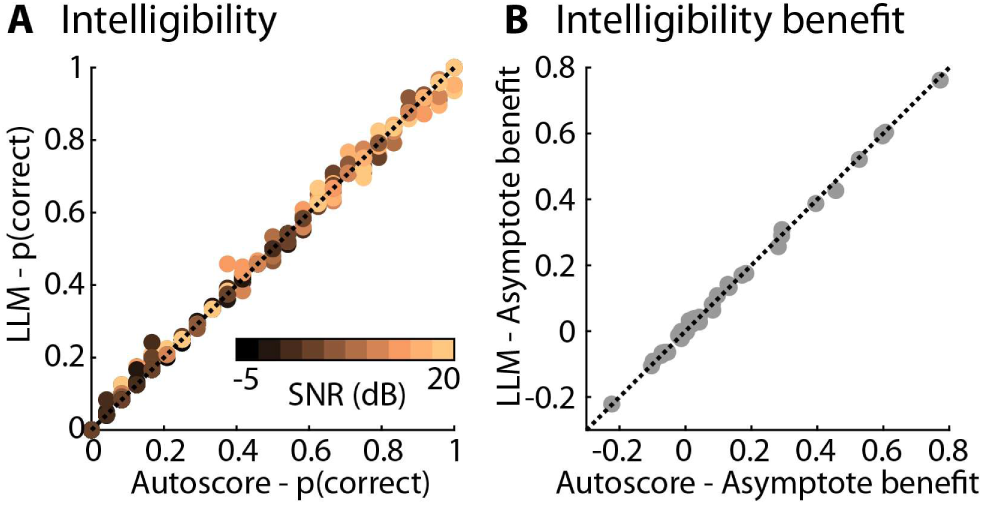
Relation between Autoscore and LLM intelligibility metrics. **A:** Relation between Autoscore and LLM for the proportion of correctly reported words per SNR, device use condition, and individual participant (color-coded). **B:** Relation between Autoscore and LLM for the hearing-aid benefit (aided minus unaided) for the estimated upper asymptote parameter from the logistic function fit, reflecting the benefit at which intelligibility plateaus. Gray dots reflect individual participants. Dotted lines indicate the identity line.

Adding PTA, audibility benefit, and age as covariates showed that PTA and audibility benefit were each independently associated with the hearing-aid benefit for the asymptote (interactions with Device Use: both F_1,35_ > 8, p < 0.008, ω^2^ > 0.06), whereas age was not (p = 0.577). The intelligibility benefit was larger for participants with worse hearing (greater PTA) and larger audibility benefit (Figure 2B, C). A greater PTA was also associated with a lower overall asymptote, that is, intelligibility plateau (F_1,35_ = 28.362, p = 6 · 10^-6^). Even when aided, participants with worse hearing understood fewer words, whereas aided intelligibility was similar for participants with smaller and larger audibility benefit (Figure 2B, C).

Finally, we quantified the agreement between LLM and Autoscore intelligibility scores. Agreement was excellent (Koo and Li, 2016), both for the proportion of correctly reported words per SNR, device use condition, and individual participant (ICC = 0.999, 95% CI [0.998–1]; Figure 3A) and for the hearing-aid benefit for the upper asymptote (ICC = 0.999, 95% CI [0.999–1]; Figure 3B). LLM scoring thus closely reproduced Autoscore, which in turn closely matches human scoring (Borrie et al., 2019; Herrmann, 2025c; Stipancic et al., 2025).

### Speech intelligibility for sentences during story listening

Analysis of the proportion of correctly reported words for a story’s final sentence revealed lower intelligibility for unaided than aided listening (Device Use: F_1,38_ = 21.364, p = 4.3 · 10^-5^, ω^2^ = 0.146) and for greater speech masking (all contrasts, t_38_ > 2.7, p_Holm_ ≤ 0.01, d > 0.23; Speech Clarity: F_2,76_ = 20.705, p = 6.6 · 10^-8^, ω^2^ = 0.097; Figure 4A). There was no difference between Autoscore and LLM scores (F_1,38_ = 0.254, p = 0.617, ω^2^ < 0.001), and there were no interactions (ps > 0.05). Scores were slightly lower overall for Gemma 4 (Figure S3).

**Figure 4:**
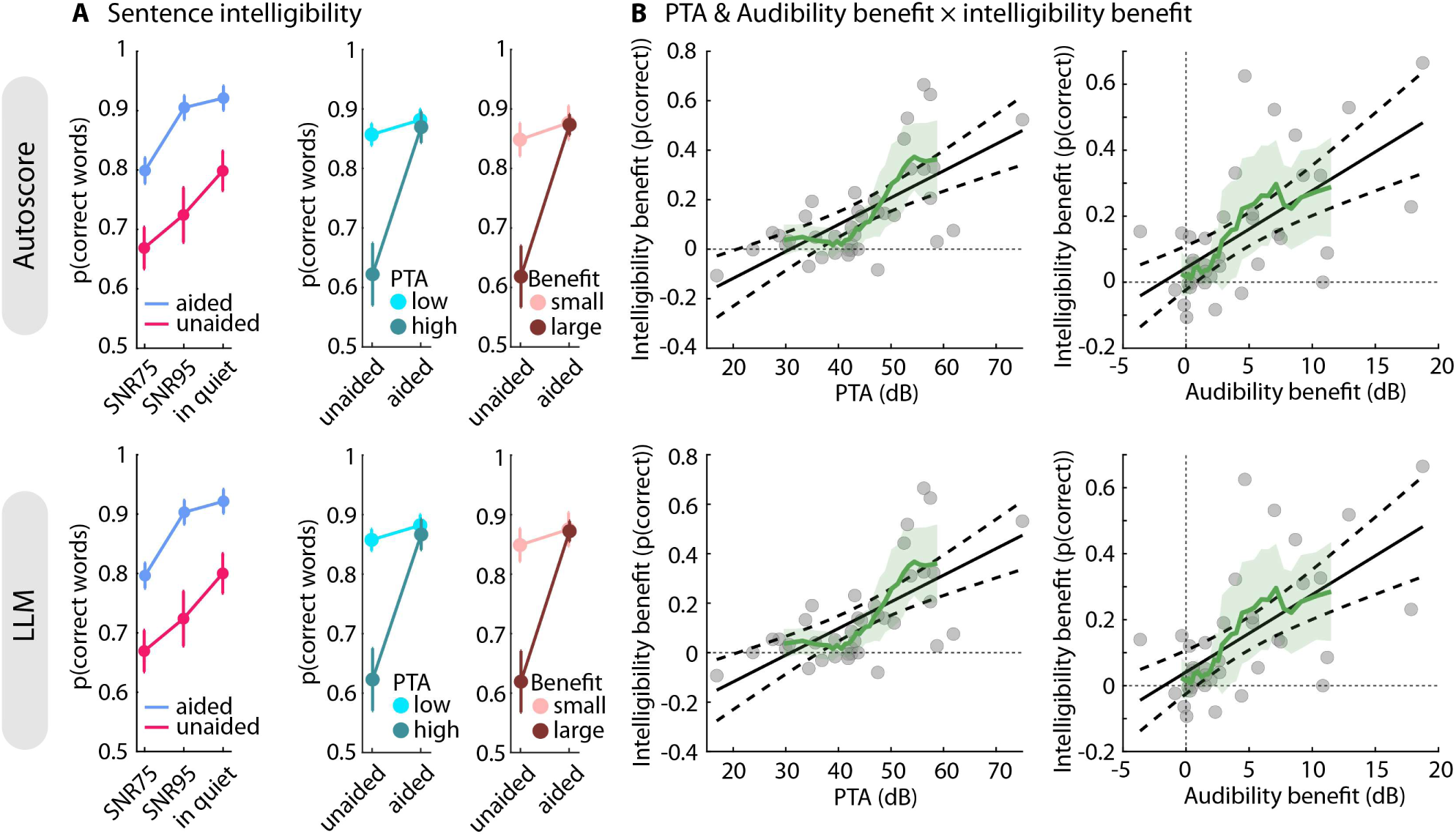
Speech intelligibility for sentences during story listening. **A:** Left: Proportion of correctly reported words of stories’ final sentence for the three speech-clarity conditions (SNR75, SNR95, in quiet), separately for aided and unaided listening. Middle and right: Speech-in-noise intelligibility averaged across speech-clarity conditions, separately for participants grouped by low (16–42.5 dB HL) vs high PTA (43.13–75 dB HL) and small (-3.63–2.32 dB) vs large audibility benefit (2.59–18.76 dB) based on median split, and separately for listening with (aided) and without hearing aids (unaided). Error bars reflect the standard error of the mean. **B:** Relationship between PTA and the intelligibility benefit (aided minus unaided; left) and between audibility benefit and the intelligibility benefit (right). Gray dots reflect individual participants. Black solid and dashed lines reflect the best linear fit and 95% confidence intervals, respectively. The green line and green shaded area reflect the mean and 95% confidence interval across 10 neighboring participants, shifted along the x-axis. The top row and bottom rows show data scored using Autoscore and an LLM, respectively.

Adding PTA, audibility benefit, and age as covariates showed that PTA and audibility benefit were independently associated with the intelligibility benefit from hearing aids (aided minus unaided; interactions with Device Use: both F_1,35_ > 17, p < 0.001, ω^2^ > 0.11), whereas age was not (p = 0.372). The intelligibility benefit increased with PTA and audibility benefit. Participants with a PTA below ∼40 dB HL or an audibility benefit below ∼2.5 dB showed little intelligibility benefit, with 95% confidence intervals including zero (Figure 4B, green line). Moreover, overall intelligibility was also lower for people with worse PTAs (F_1,35_ = 21.074, p = 5.5 · 10^-5^) and greater audibility benefits (F_1,35_ = 6.260, p = 0.017).

### Story recall: Gist and story details

Gist recall largely mirrored the sentence intelligibility effects (Figure 5), showing a lower gist recall for unaided compared to aided listening (Device Use: F_1,38_ = 12.374, p = 0.001, ω^2^ = 0.085), and lower gist recall with more speech masking (Speech Clarity: F_2,76_ = 10.865, p = 7 · 10^-5^, ω^2^ = 0.038; in quiet vs SNR75: p_Holm_ = 5.7 · 10^-4^; in quiet vs SNR95: p_Holm_ = 0.005; interaction: F_2,76_ = 0.040, p = 0.961, ω^2^ < 0.001), although recall for speech at SNR75 did not differ from recall for SNR95 (p_Holm_ = 0.075; all three contrasts were significant for Gemma 4, Figure S4). Adding PTA, audibility benefit, and age as covariates showed that PTA and audibility benefit independently predicted gist benefit from hearing aids (interactions with Device Use: both F_1,35_ >6.4, p < 0.02, ω^2^ > 0.04; Figures 5B and 5C). A better PTA was further associated with better gist recall (F_1,35_ = 15.020, p = 4.5 · 10^-4^).

**Figure 5:**
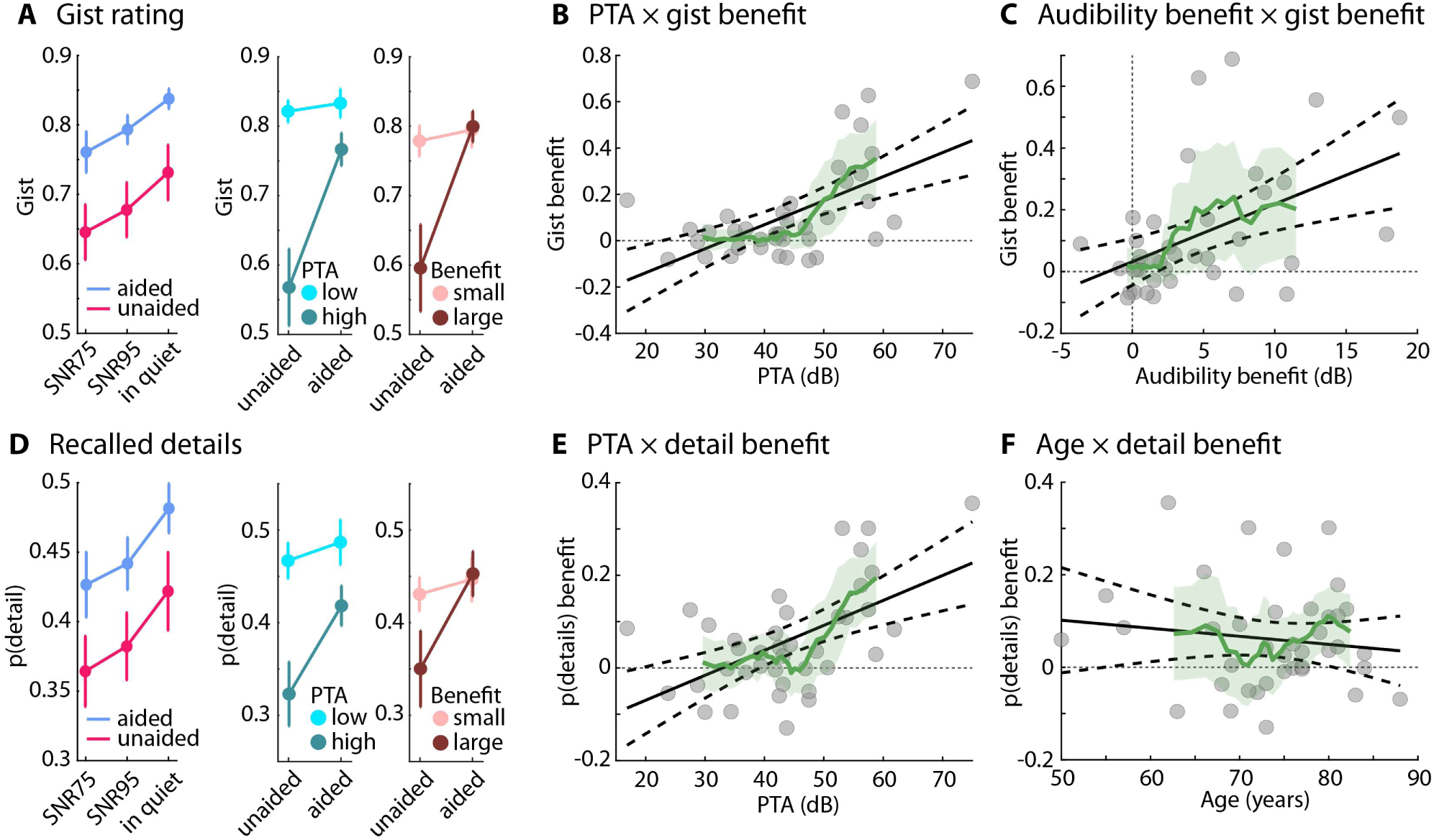
Story comprehension. **A:** Left: Gist ratings for the three speech-clarity conditions (SNR75, SNR95, in quiet), separately for aided and unaided listening. Middle and right: Gist recall averaged across speech-clarity conditions, separately for participants grouped by low (16–42.5 dB HL) vs high PTA (43.13–75 dB HL) and small (-3.63–2.32 dB) vs large audibility benefit (2.59–18.76 dB) based on median split, and separately for listening with (aided) and without hearing aids (unaided). Error bars reflect the standard error of the mean. **B:** Relationship between PTA and the gist benefit (aided minus unaided). **C:** Relationship between audibility benefit and the gist benefit. **D:** Same as A for recalled details. **E:** Relationship between PTA and the detail benefit (aided minus unaided). **F:** Relationship between age and the detail benefit. Gray dots reflect individual participants. Black solid and dashed lines reflect the best linear fit and 95% confidence intervals, respectively. The green line and green shaded area reflect the mean and 95% confidence interval across 10 neighboring participants, shifted along the x-axis.

Recall of story details was also reduced during unaided compared to aided listening (Device Use: F_1,38_ = 11.019, p = 0.002, ω^2^ = 0.049; Figure 5D), and for greater speech masking (Speech Clarity: F_2,76_ = 8.835, p = 3.5 · 10^-4^, ω^2^ = 0.031; in quiet vs SNR75: p_Holm_ = 0.002; in quiet vs SNR95: p_Holm_ = 0.014; interaction: F_2,76_ = 0.007, p = 0.993, ω^2^ < 0.001), although recalling details did not differ between SNR95 vs SNR75 (p_Holm_ = 0.190). Adding PTA, audibility benefit, and age as covariates showed independent influences of PTA and age (interactions with Device Use: both F_1,35_ > 4.4, p < 0.05, ω^2^ > 0.015; Figures 5E and 5F), but not audibility benefit (p = 0.212). A better PTA was also associated with better detail recall (F_1,35_ = 7.836, p = 0.008). The Age × Device Use interaction was not significant for the Gemma 4 model (Figure S4).

## Discussion

The present study investigated whether large language models (LLMs) can be used to assess speech perception in older adults with hearing aids, and how hearing aids affect speech intelligibility and comprehension during naturalistic story listening. LLM-based scoring closely matched established intelligibility scoring (Autoscore) for both disconnected sentences and sentences embedded in stories. On average, hearing aids improved sentence intelligibility and story recall. However, the benefit varied considerably across individuals: About half of the participants showed little or no benefit; mainly those with milder, albeit clinically relevant, hearing loss (PTA below ∼40 dB HL) and a smaller audibility benefit from their hearing aids (below ∼2.5 dB). The pattern of hearing-aid benefit was largely similar for sentence intelligibility and story recall. Contrary to our expectation that comprehension of continuous speech would be less affected than intelligibility, this suggests that hearing-aid benefits for disconnected sentences can generalize to the comprehension of continuous speech, at least for short stories at moderate SNRs assessed with free recall.

### LLM-based scoring of speech intelligibility and comprehension

Manual scoring of speech intelligibility and comprehension is labor-intensive, limiting sample sizes and the use of naturalistic speech in hearing research (Borrie et al., 2019; van Genugten and Schacter, 2024; Herrmann, 2025c). LLM text embeddings (i.e., numerical vectors representing text semantics) have been used to automate scoring by computing the semantic similarity between a response and the original speech, both for speech intelligibility (Herrmann, 2025c) and for the recall of stories and movies (Chandler et al., 2021; Heusser et al., 2021; Shen et al., 2023; Martinez, 2024; Raccah et al., 2024; Georgiou et al., 2025; Herrmann, 2025d; Panela et al., 2025). However, embeddings from language models tend to occupy a narrow region of the representational space (Ethayarajh, 2019), such that even randomly paired responses and speech materials yield non-zero similarity (Herrmann, 2025d). This makes absolute scores hard to interpret. Prompt-based LLM ratings of semantic similarity can avoid this problem (Herrmann, 2025d) but lack a direct correspondence to established metrics, especially for speech intelligibility.

For intelligibility, we thus instructed the LLM to apply the rules human scorers use, yielding the proportion of correctly reported words directly. LLM and Autoscore scores were nearly identical for individual responses and for the hearing-aid benefit (Figure 3), and statistical effects did not differ between approaches. Because Autoscore closely matches human scorers (Borrie et al., 2019; Stipancic et al., 2025), rule-based LLM scoring effectively reproduces human intelligibility scoring. It also allows context-dependent judgments (e.g., transcription variants), adaptation of scoring rules through the prompt alone, and easy generalization to other languages, for which rule-based tools and fluent human scorers are often unavailable (see also Herrmann, 2025d, for language-agnostic recall scoring).

For comprehension, we combined gist ratings with detail scoring that mirrors manual procedures. This adds to rapidly growing work using language models to score or classify free recall, including narrative recall (Georgiou et al., 2025; Panela et al., 2025), central versus peripheral details in narrative memories (Pan et al., 2025), autobiographical memories (Mistica et al., 2024; van Genugten and Schacter, 2024), and story retelling in aphasia (Kurland et al., 2025). Agreement with human raters is generally good (Chandler et al., 2021; Martinez, 2024; Mistica et al., 2024; van Genugten and Schacter, 2024; Georgiou et al., 2025; Panela et al., 2025) but can vary depending on task and implementation (Wardell et al., 2025), and LLM outputs might vary across runs, which is why we averaged scores across repeated runs. Here, we did not benchmark a specific model or scoring approach against human raters because we aimed to test whether LLM-based scoring is sensitive to speech masking and hearing-aid use in older adults. Gist and detail scores captured both effects and individual differences in hearing-aid benefit, suggesting that LLMs offer new avenues for assessing speech comprehension in hearing research and audiology. Validating detail scores against human raters remains an important next step. Finally, locally run open-source models yielded mostly comparable results (Supplementary Materials), addressing privacy concerns relevant for clinical use.

### From disconnected sentences to story comprehension

Hearing-aid benefit is typically assessed with disconnected words or sentences, yet intelligibility only weakly predicts comprehension, particularly at poor SNRs (Fontan et al., 2015). Age-related deficits seen for isolated sentences in unfavorable conditions can be reduced or absent for discourse and stories (Tye-Murray et al., 2008; Irsik et al., 2022), and older adults can comprehend spoken passages in noise as well as younger adults when listening conditions are adjusted for hearing (Schneider et al., 2000). We thus expected gist recall to be less affected than intelligibility by masking and hearing-aid use. Contrary to this expectation, gist recall and recall of details largely tracked intelligibility. All were lower for unaided than for aided listening and lower at SNR75 and SNR95 than in quiet, and the speech intelligibility and recall benefit was predicted by PTA and audibility benefit. The only difference emerged for SNR75 vs. SNR95, where intelligibility, but not gist and detail recall, differed. Detail recall behaved somewhat differently from gist recall, because its hearing-aid benefit was associated with age but not with audibility benefit.

Three aspects of the current design may have contributed to why gist and detail recall mirrored intelligibility. First, the SNRs were moderate. Mean SNRs were 8.8 dB (SNR75) and 18.5 dB (SNR95), comparable to everyday listening, where SNRs are mostly positive (Smeds et al., 2015; Wu et al., 2018). Most words are understood at these SNRs, whereas intelligibility and comprehension might diverge more at poorer SNRs (Fontan et al., 2015). Second, the stories were short (∼120 words). The gist of a short story rests on only a few key ideas, so there is little redundancy to compensate for missed words. Third, the studies showing robust story comprehension assessed it with questions (Schneider et al., 2000; Tye-Murray et al., 2008), which cue the relevant content and can often be answered from partial understanding or inference, whereas free recall provides no such cues. Detail recall, in turn, may have been reduced because understanding speech in noise takes effort even when most words are understood, which leaves less capacity for storing what was heard (McCoy et al., 2005). Consistent with this, acoustic degradation reduces memory for spoken stories even when nearly all words are understood (Ward et al., 2016). That the hearing-aid benefit for detail recall was related to age but not to audibility benefit likewise suggests that factors other than audibility contribute.

The current study cannot answer whether hearing-aid benefits may differ between intelligibility and gist comprehension for different overall speech levels, lower SNRs, longer materials, delayed recall, or conversations, in which context accumulates and motivation and fatigue shape listening effort. Nevertheless, the LLM-based approach makes such extensions feasible, because stories of any length can be generated and scored automatically.

### Who benefits from hearing aids?

On average, hearing aids improved sentence intelligibility and story recall, but the benefit varied widely across individuals. It increased with PTA and audibility benefit, but participants with a PTA below ∼40 dB HL or an audibility benefit below ∼2.5 dB showed little benefit (Figures 2, 4, and 5). Here, this was the case for about half of the participants, even though all but two had clinically significant hearing loss (PTA >25 dB HL). That hearing loss partially predicts aided speech recognition in older adults is well established (Humes, 2002). Moreover, previous work shows – consistent with our work – that the hearing-aid benefit for soft speech is greatest for listeners with a PTA above 40 dB HL (Shanks et al., 2002) and that self-reported benefit is lowest for mild hearing loss (Ziemska-Gorczyca et al., 2024). However, some participants with poorer hearing also did not benefit (Figures 4 and 5), indicating that hearing-aid benefit can differ between listeners with similar thresholds. We show that audibility benefit predicted the hearing-aid benefit independently of PTA, suggesting that how much audibility an individual’s hearing aids provide matters in addition to the degree of hearing loss. Indeed, aided speech perception is poorer when hearing-aid output falls below prescription targets (Engler et al., 2026), and fittings may deviate from these targets unless verified with real-ear measurements (Aazh and Moore, 2007).

Hearing-aid benefit also depended on the SNR (Figure 2). For disconnected sentences, hearing aids raised the upper asymptote of the intelligibility function but did not change its slope or midpoint, so the benefit was largest at favorable SNRs, where intelligibility plateaus. The stories were presented at such SNRs (mean: 8.8 and 18.5 dB), which are somewhat more favorable than the about +5 dB typical of everyday listening in noise (Smeds et al., 2015; Wu et al., 2018). In noisier settings, the benefit is thus likely smaller, and the proportion of users with little benefit may be larger, although differences between intelligibility and gist recall may emerge. Nevertheless, participants wore their hearing aids for a median of 11 hours per day and were satisfied with them, consistent with large surveys (Picou, 2020, 2022), and those with a larger audibility benefit wore them longer each day. Hearing aids may thus provide benefits not captured by the present measures, for example for soft speech, awareness of environmental sounds, or listening effort. Clinically, this may suggest counseling hearing-aid candidates, particularly those with milder hearing loss, that improvements in understanding speech may be modest, especially in noise, while other benefits may still be meaningful.

## Conclusion

The current study shows that LLM-based scoring provides an automated approach to assessing speech intelligibility and story comprehension in older hearing-aid users. Hearing-aid benefit for story recall was comparable to that for sentence intelligibility, at least for short stories presented at favorable SNRs. Benefit increased with hearing loss and audibility benefit, and was small for about half of the participants, mainly those with milder hearing loss. LLM-based assessment of continuous speech may help determine for whom and under which listening conditions hearing aids improve speech comprehension.

## Supporting information

Supplementary Materials and Results

## Data availability

Data will be available upon reasonable request to the corresponding author.

## Author Contributions

**YR:** Conceptualization, methodology, investigation, data curation, writing - review & editing. **NI:** Investigation, data curation, writing - review & editing. **SB:** Investigation, data curation, writing - review & editing. **TR:** Methodology, investigation, writing - review & editing. **SC:** Conceptualization, methodology, investigation, writing - review & editing. **KAG:** Conceptualization, methodology, investigation, writing - review & editing. **BH:** Conceptualization, methodology, validation, formal analysis, investigation, data curation, writing - original draft, writing - review & editing, visualization, supervision, project management, funding acquisition.

## Acknowledgements

The research was supported by the Canada Research Chair program (CRC-2023-00383), the William Demant Foundation, and the Data Science Institute of the University of Toronto. Language editing (grammar correction, sentence shortening, and paragraph condensing) was assisted by Claude Opus 5.5 (Anthropic; https://claude.ai). The authors reviewed all revisions and take full responsibility for the final text.

## Statements and Declarations

The authors have no conflicts or competing interests.

