## Supplementary Materials and Results for "Examining hearing-aid benefit during story listening in older adults using large language models"

### Supplementary methods and materials

#### LLM prompts for story generation and data analysis

This section provides the exact prompts used for story generation and for LLM-based scoring of speech intelligibility, story gist, and story details. The same prompts were used for the proprietary (OpenAI) and the open-source models described in the main text. In the prompts, text in curly brackets (e.g., {topic}) denotes placeholders that were replaced by the respective content for each call. All LLM-based analyses used a temperature of 1 and were repeated five times per response, and scores were averaged across repetitions.

##### *Story generation*

OpenAI's GPT-4o ('gpt-4o-2024-08-06'; OpenAI et al., 2023) was used to generate 18 story texts (Georgiou et al., 2025; Herrmann, 2025a). Each story text focused on a different topic (e.g., a rivalry that turns into respect). The model was prompted using a zero-shot approach (Chen et al., 2025) with the following input:

> *"Write an engaging story about {topic}. The story should be exactly 120 words long and use simple vocabulary. The story should be written in a single, continuous paragraph without any line breaks. Focus on building a rich context where each detail adds to a deeper understanding of the main plot. The final sentence should contain exactly 10 words, summarizing or concluding the story in a way that only makes sense if the reader has absorbed the entire context. Make sure the narrative gradually reveals important information so that understanding the ending strongly depends on the context provided throughout."*

Stories were manually verified and minimally adjusted to ensure coherence. We requested 10 words for the final sentence because the model often produced fewer words than requested, and we targeted 8 words as in the sentence-in-noise task. Final sentences that felt detached from the story or contained unwanted symbols were regenerated using the following prompt:

> *"Write a final sentence for this story: {story}. The final sentence should contain exactly 8 words, summarizing or concluding the story in a way that only makes sense if the reader has absorbed the entire context. Do not include any specific names or unique identifiers. Avoid using commas, colons, or semicolons."*

The final sentences of the stories used in the study comprised eight words, except for two stories whose final sentences comprised seven words.

##### *Speech intelligibility scoring*

In addition to Autoscore (Borrie et al., 2019), speech intelligibility was scored using OpenAI's 'gpt-5-mini-2025-08-07' model, with scoring rules implemented in the prompt that follow those of Autoscore (http://autoscore.usu.edu/). Before scoring, target sentences and response transcripts were Unicode-normalized, typographic apostrophes were replaced by the standard apostrophe, texts were converted to lower case, and words were extracted, removing punctuation and apostrophes at word edges. The words of the target sentence and of the response transcript were then provided to the model as numbered lists, with each word in quotation marks (e.g., 1. "rescue" 2. "arrived" …). The system message was 'You are an expert scorer of speech intelligibility.', and the prompt was:

> *Act as an expert scorer of speech intelligibility for a research study. A participant heard a target sentence and reported aloud what they heard; their response was transcribed. Both have been split into numbered words below.*
>
> *TARGET WORDS:*
> *{target word list}*
>
> *TRANSCRIPT WORDS:*
> *{transcript word list}*
>
> *Go through the target words one by one and decide whether the participant reported each one, using these rules:*
> *1. Position and word order do not matter. A target word may be matched by a transcript word at any position.*
> *2. Each transcript word may be used for at most one target word.*
> *3. Judge by pronunciation, not spelling: homophones and plausible transcription spellings count as matches (e.g. 'their' for 'there', 'nite' for 'night').*
> *4. Morphological changes do NOT match: differences in number, tense, or affixes (e.g. 'dog' vs 'dogs', 'walk' vs 'walked', 'is' vs 'was') count as not reported.*
> *5. Synonyms and paraphrases do NOT match. Meaning is irrelevant; only the word matters.*
> *6. Unmatched transcript words are ignored and never penalised.*
> *7. Each numbered entry is exactly one word, even if it contains an apostrophe. Never split an entry into two words.*
> *8. Each target word is either reported or not reported. There is no partial credit: never award a fraction of a word.*
>
> *Then respond with the number of target words reported correctly, followed by a forward slash, followed by the total number of target words. Both must be whole numbers; decimals are not valid. Do not count the target words yourself: the total is simply the number of the last entry in the TARGET WORDS list above. Output only the score, for example 4/8. No other text.*

A response was accepted if it consisted of two whole numbers separated by a forward slash, the total matched the actual number of target words, and the number of correctly reported words did not exceed the total. Otherwise, the prompt was resubmitted (maximum of three attempts). The proportion of correctly reported words was calculated as the number of correctly reported words divided by the number of target words. Missing responses were submitted as empty transcripts. The numbers of correctly reported words and of target words were each averaged across the five repetitions.

***Gist rating of story recall***

The gist of participants' story recall was rated using OpenAI's 'gpt-5-mini-2025-08-07' model, following previous work (Herrmann, 2025b; Panela et al., 2025). The model was provided with the original story text and the participant's recall transcript. The system message was 'You are an expert rater of speech contents.', and the prompt was (reproduced verbatim):

> *Act as an expert rater of speech contents. For a research study, we obtained a participant's recall of a short story. I would like you to rate the degree to which the gist of the story was recalled by the participant. On a scale from 0-100 (in steps of 1), rate the degree to which the recall transciptfrom the participant captures the story text. 0 refers to "not very" and 100 refers to "very". If the recall transcipt mostly does not capture the gist of the story text, give a 0. If the recall transcipt mostly captures the gist of the story text, give a 100. Use the full range of the scale. That is, do not hesitate giving a 0 or 100 if applicable. This is the story text: {story text}". This is the recall transcript: {recall transcript}". Please provide only the rating score and nothing else.*

A response was accepted if it consisted of a whole number; otherwise, the prompt was resubmitted (maximum of three attempts). Ratings were divided by 100, yielding gist scores between 0 and 1. Ratings were averaged across the five repetitions.

***Detail scoring of story recall***

Recall of story details was scored in two steps using OpenAI's 'gpt-5-mini-2025-08-07' model. In the first step, each original story text was decomposed into a list of atomic recallable details, which served as the scoring key for the recall transcript. The definition of atomic details follows the notion of propositions (Kintsch and van Dijk, 1978), idea units (Sisco et al., 2013), details (Sacripante et al., 2023; van Genugten and Schacter, 2024), or clauses (Shen et al., 2023; Georgiou et al., 2025) used to score text and narrative recall. The system message was 'You are an expert analyst for memory and discourse research. You break text from stories or recall into atomic recallable details.', and the prompt was:

> *Your task is to break the text into ATOMIC RECALLABLE DETAILS.*
>
> *Definition:*
> *- An atomic detail is a single fact, event, state, relationship, or outcome that a participant could recall independently.*
> *- Each detail must express only ONE idea.*
> *- Do not merge multiple facts into one detail.*
> *- Include explicit details from the text.*
> *- Include central causal or relational details only if clearly stated.*
> *- Do NOT include speculative inferences.*
> *- Keep wording simple and neutral.*
> *- Prefer concrete facts.*
> *- Theme or moral details are allowed only if explicitly stated in the text.*

*Return ONLY valid JSON in this exact format:*
*{*
*"details": [*
*"...",*
*"...",*
*"..."*
*]*
*}*

*Text:*
*"""{story text}"""*

The output was constrained to a JSON object containing a single list of text strings (structured output with a JSON schema). Empty entries were removed, and the prompt was resubmitted if the output could not be parsed (maximum of three attempts).

In the second step, each recall transcript was scored against the details of the corresponding story, which were provided to the model as a numbered list (e.g., 1. {detail 1} 2. {detail 2} …). The system message was 'You are an expert scorer for a memory experiment. You compare participant recall against a fixed set of atomic story details.', and the prompt was:

*Compare the participant's recall to the list of atomic story details.*

*For each detail, assign exactly one score:*
*- 1 = recalled*
*- 0.5 = partly recalled*
*- 0 = not recalled*

*Instructions:*
*- Score based on meaning, not exact wording.*
*- Accept paraphrases.*
*- Be liberal rather than conservative in assigning scores.*
*- If the recall reasonably captures the meaning of a detail, give credit.*
*- Use 1 when the essential content of the detail is present.*
*- Use 0.5 when the gist is present but important specificity is missing.*
*- Use 0 when the detail is absent, too vague, or unsupported.*

*Return ONLY a Python list of numbers.*
*- No explanation*
*- No labels*
*- No JSON*
*- No markdown*

*- The list must have exactly {number of details} values*
*- Keep the same order as the detail list below*

*Atomic story details:*
*{numbered detail list}*

*Participant free recall:*
*"""{recall transcript}"""*

A response was accepted if it contained a list with exactly one score per detail and each score was 0, 0.5, or 1; otherwise, the prompt was resubmitted (maximum of three attempts). The three-level scoring follows manual story-recall scoring with partial credit (Power et al., 1979; Sisco et al., 2013; Martinez, 2024). The proportion of recalled details was calculated as the sum of scores divided by the number of details. Both steps used a temperature of 1 and were repeated five times for each recall transcript (with the story details extracted anew in each repetition), and the proportion of recalled details was averaged across the five repetitions.

#### Supplementary results

Supplementary results are provided for data analyzed using open-source, locally run LLMs. Alibaba's Qwen3-ASR-1.7B was used for transcription and Google's Gemma 4 ('gemma-4-26b-a4b', executed using 4-bit quantized GGUF weights) for the analysis of speech intelligibility, gist recall, and detail recall scoring.

##### Intelligibility of disconnected sentences in multi-talker babble

For the sentence-in-noise task, the upper asymptote, reflecting the performance plateau at high SNRs, was higher for aided than unaided listening (Device Use: $F_{1,38} = 19.588$, $p = 7.8 \cdot 10^{-5}$, $\omega^2 = 0.111$; Figure S1) and did not differ between Autoscore and LLM scoring (Analysis Approach: $F_{1,38} = 2.456$, $p = 0.125$, $\omega^2 < 0.001$; interaction: $p > 0.15$; Figures S1 and S2). The midpoint of the logistic function fit was greater for Gemma 4 than Autoscore ($F_{1,38} = 18.354$, $p = 1.2 \cdot 10^{-4}$, $\omega^2 = 0.013$). Other effects or interactions for the slope and midpoint were not significant (all ps > 0.12).

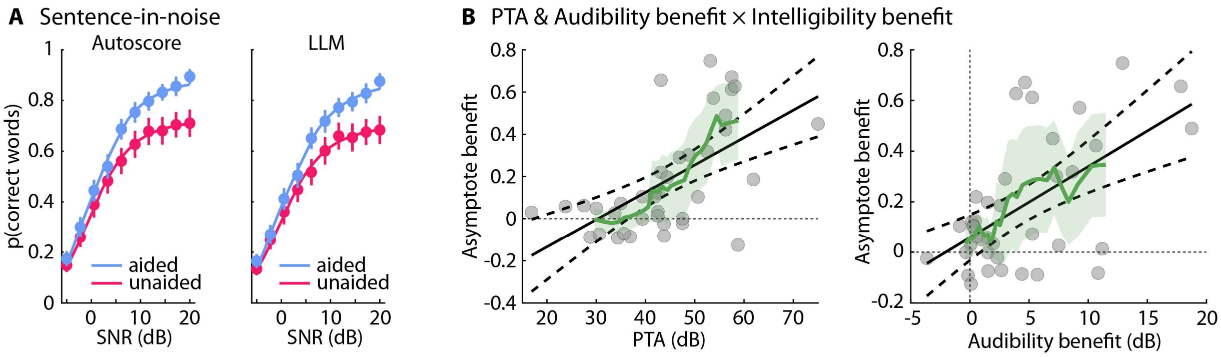

**Figure S1: Speech intelligibility for disconnected sentences. A:** Proportion of correctly reported words for Autoscore and the LLM (Gemma 4). **B:** Relationship between PTA and the intelligibility benefit (i.e., upper asymptote) and the relationship between audibility benefit and the intelligibility benefit (aided minus unaided) for Gemma 4. PTA and audibility benefit both contributed independently to the intelligibility benefit. Gray dots reflect individual participants. Black lines reflect the best linear fit and 95% confidence intervals. The green line and green shaded area reflect the mean and 95% confidence interval across 10 neighboring participants, shifted along the x-axis.

Adding PTA, audibility benefit, and age as covariates showed that PTA and audibility benefit were each independently associated with the hearing-aid benefit for the asymptote (interactions with Device Use: both $F_{1,35} > 10$, $p < 0.005$, $\omega^2 > 0.065$), whereas age was not ($p = 0.820$). The intelligibility benefit was larger for participants with worse hearing (greater PTA) and larger audibility benefit (Figure S1B). A greater PTA was also associated with a lower overall asymptote, that is, intelligibility plateau ($F_{1,35} = 30.148$, $p = 3.6 \cdot 10^{-6}$). Even when aided, participants with worse hearing understood fewer words, whereas aided intelligibility was similar for participants with smaller and larger audibility benefit.

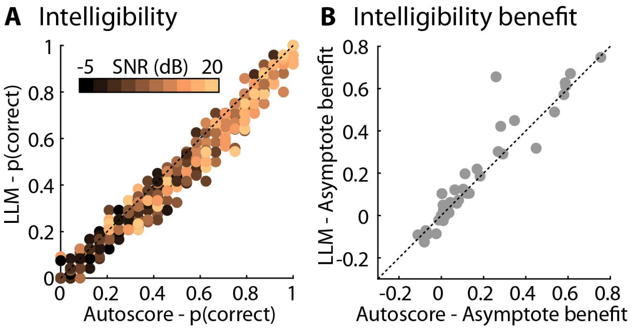


**Figure S2: Relation between Autoscore and LLM intelligibility metrics. A:** Relation between Autoscore and LLM for the proportion of correctly reported words per SNR, device used condition, and individual participant (color-coded). **B:** Relation between Autoscore and LLM for the hearing-aid benefit (aided minus unaided) for the estimated upper asymptote parameter from the logistic function fit, reflecting the benefit at which intelligibility plateaus. Gray dots reflect individual participants. Dotted lines indicate the identity line.

We quantified the agreement between LLM and Autoscore intelligibility scores. Agreement was excellent (Koo and Li, 2016), both for the proportion of correctly reported words per SNR, device use condition, and individual participant (ICC = 0.988, 95% CI [0.968, 0.994]; Figure S2A) and for the hearing-aid benefit for the upper asymptote (ICC = 0.946, 95% CI [0.899, 0.971]; Figure S2B).

**Sentence intelligibility for sentences during story listening**

Analysis of the proportion of correctly reported words for a story's final sentence revealed lower intelligibility for unaided than aided listening (Device Use: $F_{1,38} = 22.404$, $p = 3 \cdot 10^{-5}$, $\omega^2 = 0.139$) and for greater speech masking (all contrasts, $t_{38} > 2.5$, $p_{Holm} \leq 0.015$, $d > 0.22$; Speech Clarity: $F_{2,76} = 22.831$, $p = 1.7 \cdot 10^{-8}$, $\omega^2 = 0.093$; Figure S3A). Scores from Gemma were lower than those for Autoscore ($F_{1,38} = 99.871$, $p = 3.5 \cdot 10^{-12}$, $\omega^2 < 0.013$), and there was additional nuance related to speech clarity (Approach × Speech Clarity interaction: $F_{2,76} = 7.789$, $p = 8.4 \cdot 10^{-4}$, $\omega^2 = 0.001$).

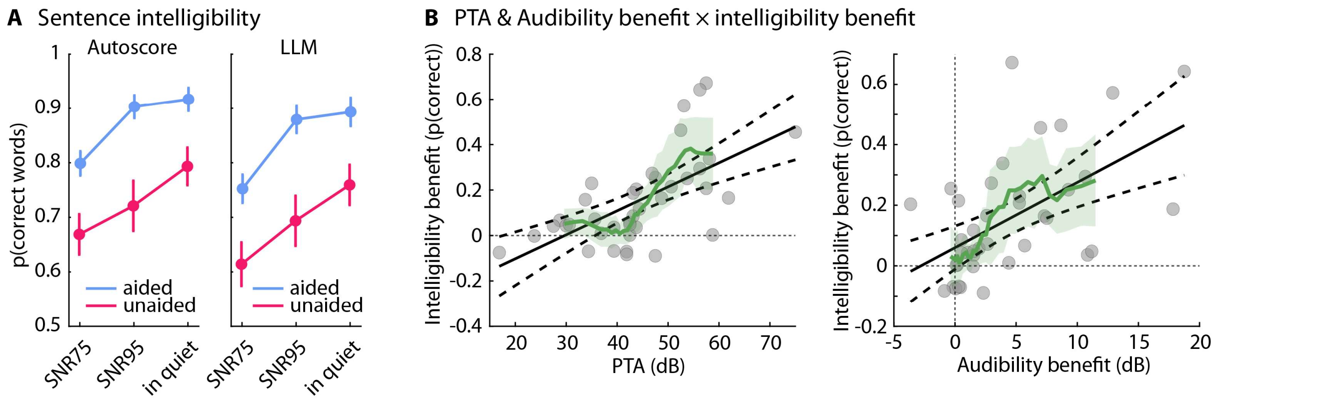


**Figure S3: Speech intelligibility for sentences during story listening. A:** Proportion of correctly reported words of stories' final sentence for the three speech-clarity conditions (SNR75, SNR95, in quiet), separately for aided and unaided listening, and separately for Autoscore and the LLM (Gemma 4). Error bars reflect the standard error of the mean. **B:** Relationship between PTA and the intelligibility benefit (aided minus unaided; left) and between audibility benefit and the intelligibility benefit (right) for Gemma 4. Gray dots reflect individual participants. Black solid and dashed lines reflect the best linear fit and 95% confidence intervals, respectively. The green line and green shaded area reflect the mean and 95% confidence interval across 10 neighboring participants, shifted along the x-axis.

Adding PTA, audibility benefit, and age as covariates showed that PTA and audibility benefit were independently associated with the intelligibility benefit from hearing aids (aided minus unaided; interactions with Device Use: both $F_{1,35} > 13$, $p < 0.001$, $\omega^2 > 0.08$), whereas age was not ($p = 0.284$). The intelligibility benefit increased with PTA and audibility benefit. Participants with a PTA below ~40 dB HL or an audibility benefit below ~2.5 dB showed little intelligibility benefit, with 95% confidence intervals including zero (Figure S3B, green line). Moreover, overall intelligibility was also lower for people with worse PTAs ($F_{1,35} = 19.846$, $p = 8.2 \cdot 10^{-5}$) and greater audibility benefits ($F_{1,35} = 7.682$, $p = 0.009$).

#### Story recall: Gist and story details

Gist recall was lower for unaided compared to aided listening (Device Use: $F_{1,38} = 15.894$, $p = 2.9 \cdot 10^{-4}$, $\omega^2 = 0.115$; Figure S4A), and lower with more speech masking (for all contrasts: $t_{38} > 2.09$, $p_{Holm} < 0.05$; Speech Clarity: $F_{2,76} = 13.580$, $p = 9 \cdot 10^{-6}$, $\omega^2 = 0.061$; interaction: $F_{2,76} = 1.130$, $p = 0.328$, $\omega^2 < 0.001$). Adding PTA, audibility benefit, and age as covariates showed that PTA and audibility benefit independently predicted gist benefit from hearing aids (interactions with Device Use: both $F_{1,35} > 12$, $p < 0.002$, $\omega^2 > 0.08$; Figures S4B and S4C). A better PTA ($F_{1,35} = 19.905$, $p = 8.1 \cdot 10^{-5}$) and a lower audibility benefit ($F_{1,35} = 6.784$, $p = 0.013$) were further associated with better overall gist recall.

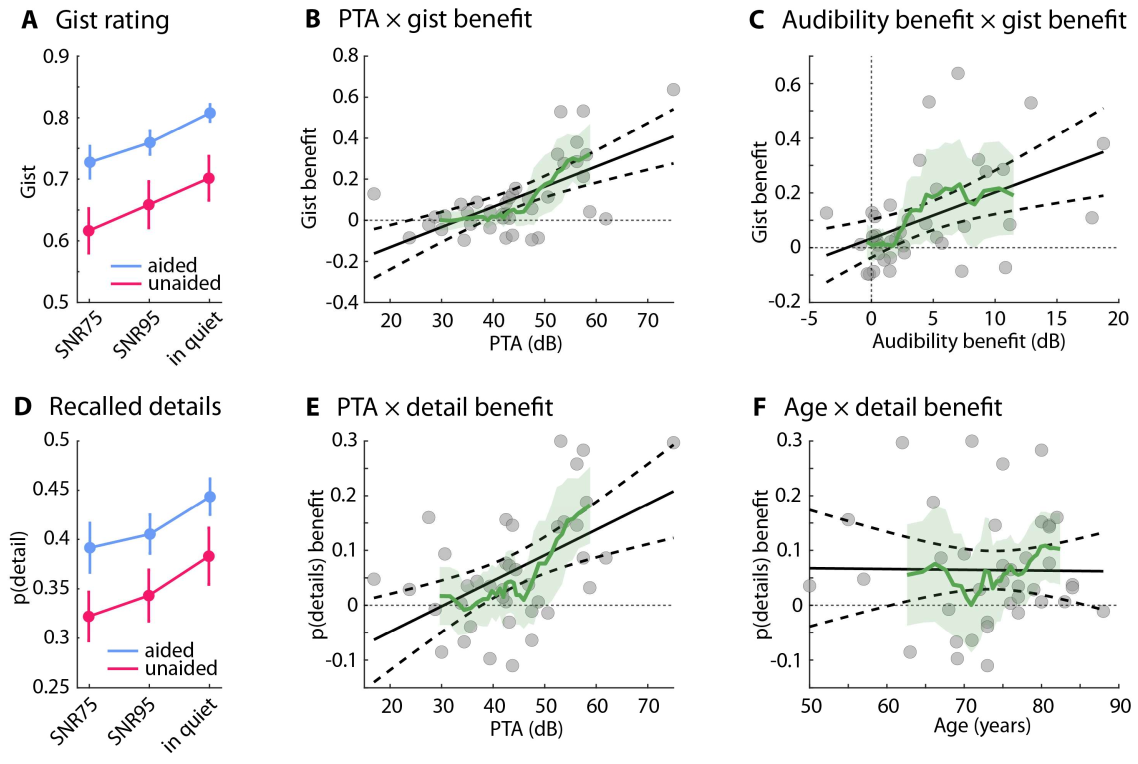


**Figure S4: Story comprehension. A:** Gist ratings for the three speech-clarity conditions (SNR75, SNR95, in quiet), separately for aided and unaided listening. Error bars reflect the standard error of the mean. **B:** Relationship between PTA and the gist benefit (aided minus unaided). **C:** Relationship between audibility benefit and the gist benefit. **D:** Proportion of recalled story details, separately for aided and unaided listening. **E:** Relationship between PTA and the detail benefit (aided minus unaided). **F:** Relationship between age and the detail benefit. Gray dots reflect individual participants. Black solid and dashed lines reflect the best linear fit and 95% confidence intervals, respectively. The green line and green shaded area reflect the mean and 95% confidence interval across 10 neighboring participants, shifted along the x-axis.

Recall of story details was also reduced during unaided compared to aided listening (Device Use: $F_{1,38} = 14.296$, $p = 5.4 \cdot 10^{-4}$, $\omega^2 = 0.052$; Figure S4D), and for greater speech masking (Speech Clarity: $F_{2,76} = 9.802$, $p = 1.6 \cdot 10^{-4}$, $\omega^2 = 0.029$; in quiet vs SNR75: $p_{Holm} = 7.4 \cdot 10^{-4}$; in quiet vs SNR95: $p_{Holm} = 0.006$; interaction: $F_{2,76} = 0.055$, $p = 0.964$, $\omega^2 < 0.001$), although recalling details did not differ between SNR95 vs SNR75 ($p_{Holm} = 0.179$). Adding PTA, audibility benefit, and age as covariates showed that hearing-aid benefit for the recall of details was greater for people with a greater PTA (interaction with Device Use: $F_{1,35} = 10.293$, $p = 0.003$, $\omega^2 = 0.037$; Figure S4E), whereas audibility benefit and age did not predict the benefit from hearing aids for the recall of details. A better PTA was also associated with better detail recall ($F_{1,35} = 6.317$, $p = 0.017$).
